# Additive Compendium Map of Outbreak Risk Determinants of West Nile Virus in Europe at NUTS3

**DOI:** 10.1101/2020.08.27.20183194

**Authors:** Alberto Alexander Gayle

## Abstract

Annual emergence of West Nile virus depends on a complex transmission chain. Predictive efforts are consequently confounded by time-varying associations and scale-dependent effect variability. SHAP (SHaply Additive Explanation) is a novel AI-driven solution with potential to overcome this. SHAP takes a high-performance XGBoost model and deductively imputes the marginal contribution of each feature with respect to the log relative risk associated with the local XGBoost prediction (an additive model). The resulting effect matrix is dimensionally identical to the original data but IID and homogenized in terms of units, scale, and interpretation. Such “synthetic data” can therefore serve as surrogate to allow for high-power statistical analyses. Here, we applied SHAP to a database consisting of high-resolution data from various domains – climate, environment, economic, sociodemographic, vector and host distribution – to derive an effect matrix of WNV outbreak risk determinants in Europe. This effect data proved superior to the original, nominal data in predictive tasks and delivered qualitatively compelling, domain-specific risk mappings. Further applications are discussed and others are invited to experiment.

## Background & Summary

West Nile Virus (WNV) is a mosquito-borne virus with deadly potential^1^. The virus has since rapidly spread in recent years and now enjoys the widest geographic distribution of any similar mosquito-borne viruse^2^. 2018 alone saw a 7.2 fold increase in the number of cases reported in Europe along with a markedly expanded geographic range (see *Figure* 1). Many factors have been associated with this spread, but much is still unknown^3^. The endemic potential of WNV is not to be underestimated: seropositivity is reportedly as high as 90% in the sub-Saharan regions where WNV has long been endemic^4^. Reports of human-to-human transmission routes – via blood transfusion^5^, organ transplants^6^, aerosol^7,8^, birthing and breast feeding^9^, and possibly even sexual contact^10^- have added to the concern^11,12^. Response options are limited to symptomatic treatment and no human vaccine is available^10^. And the continued emergence of multiple lineages^13-16^, each with distinct transmission and syndromatic profiles^17^, adds to the uncertainties. Predicting annual emergence and spread of WNV in Europe has therefore become a priority.

The spread of West Nile virus is dependent on a wide range of determinants, including climatic features, environment, and sociodemographic factors^18^. However, predictive modeling efforts have often been confounded by complex interactions and time-varying associations^19^. Early warning models can therefore often be difficult to implement in practice^20^. Various analytical modalities have been used for this purpose, each with different pros and cons. Most have met with varying degrees of success, but scope- or context-dependent inconsistency of effect remains an outstanding issue^21^. Classification tree models are a promising entry in this field^22^. However, some controversy has been merited. Most notably, questions persist as to the long-term predictive accuracy of these models. This is due to the “black box” nature of these models whereby the mechanistic structure being assumed by these models are not open to scrutiny: unlike traditional models, no parameter estimates are provided^22^. This makes such models particularly unsuited for the early warning case, or intervention strategy, as 1) direct effect of specific predictors on the predictive outcomes cannot be determined, and 2) there is no way to control or investigate the reliability of the deterministic structure driving predictions.

SHAP (SHaply Additive Explanations)^23^ is a novel AI-driven solution with potential to overcome overcome the problems associated with classification tree models. SHAP takes a high-performance XGBoost model^24^ and deductively imputes feature effects and predictive structure on a case-wise basis^25^. The effect matrix derived from the resulting SHAP meta-model is dimensionally identical to the original data set and can be summarized to produce effect estimates and analyses particular to any given subset of cases or features. The underlying predictive engine, XGBoost, is notably robust against multicollinearity^26^, automatically facilitates data curation^27^, and outperforms even the most state-of-the-art prediction methods for most types of structured data^28^. SHAP has been mathematically proven to be optimal with respect to computing additive contribution (effects) of individual features to a given event^29^. Provided sufficiently high performancing predictive model, the determinant substructure revealed by SHAP can therefore be assumed to enjoy a high-degree of empirical validity – more so provided a sufficiently challenging validation task.

Herein, we report the application of the XGBoost-SHAP pipeline to generate to a large, high-resolution data set consisting of marginal feature effects predictive of WNV outbreak risk. This data set is comprehensive with respect to a broad set of knowledge domains, including climate, environment, economic, sociodemographic, vector distribution, and host presence. The extraordinary WNV outbreak of 2018 in Europe was selected to validate the original predictive model due to the extreme degree to which it differed from prior years: high out- of-sample performance would imply an empirically valid model. Empirical validity was assessed by comparing the performance of SHAP-derived effect matrix to the original, nominal data with respect to traditional multivariate analyses. SHAP was found to perform better than the original, nominal data at all levels of model complexity for all feature classes. The Shapley heuristic allows for individually meaningful domain-specific feature effect aggregations. We therefore evaluated this property as well with respect to linear regression. Here too we found that the effect data produced by SHAP performed well, even when applied to predict alternative outputs. The risk mappings associated with each aggregate were furthermore found to be statistically independent and qualitatively valid with respect to each respective domain. Several high-power analytical applications are presented in a forthcoming manuscript, available as a pre-print online (Gayle 2020a)^54^. The geotagged, SHAP-derived effect data is provided for Europe along with the corresponding nominal data.

## Methods

### Original data collection

A literature review was conducted. The purpose of this review was to identify potential covariates of West Nile virus (WNV) emergence. PubMed and Google Scholar were searched. All research published between 1999 and 2019 was identified that included “West Nile” in either the title or abstract. This set of research publications was furthermore filtered. Only those titles or abstracts that referred to geospatial risk associations were included. Search terms for this secondary filter included: “risk”, “model”, “predict”, “estimat”, “associat”, “determinant”, “factor” and “review”. This list was then manually screened to exclude articles that presented neither primary research nor references to the results of such. The determinants presented were then extracted and compiled. Time-lag and other interaction effects were noted separately and in addition to the primary determinants. Determinant specifications with respect to aggregation, normalization, and other processing were also noted. This list was then stratified into eight categories based on class and source of data. Spatial and temporal covariates were included among the eight feature classes.

The selected feature classes included the following broad categories: “climate”, “environmental”, “economic”, “sociodemographic”, “hosts”, and “vectors”. Public data sources for each feature class were identified and selected based on the availability of data that aligned with determinant specifications. The literature on WNV consistently describes it as “hard to predict”. And this is reflected in broad uncertainties surrounding the precise mechanistic associations surrounding even the most basic, and generally accepted, determinants. In many cases, time-lagged climatic features are included in models to serve as proxy for specific features relevant to vector breeding or disease transmission. For example, what does it mean to associate rain that occurred 4 weeks prior to a reported case today? What is the actual biotic or environmental feature that is driving increased risk today: a) precipitation accumulation, b) soil moisture, c) standing water, or d) increased surface reflectivity indicative of water presence (MNDWI)? In order to avoid such potentially confounding effects, we expanded our inclusion criteria to include all climatic or environmental indicators available from a given selected source – this included some features not present in the literature.

All data was downloaded and processed using the R platform. Spatial and temporal covariates were included: one for each of the district-level regions in Europe (NUT3) as well as years 2010-2016 and 2018. Geometries for each of the NUTS3 regions were obtained using the “Eurostats” package in R. Public sources were sought out for each of the remaining feature classes. The “earthEngineGrabR”^30^ package in R was used to obtain climatic data via the Google Earth Engine: this included the highly-cited TerraClimate^31^, MerraClim^32^, Global Surface Water^33^ data sets. Climatic trend and anomaly data was obtained directly, in preprocessed form from the International Research Institute for Climate and Society (IRI) at Columbia University (New York, USA)^34^. The “Bioclimatic” feature set is a well-cited set of time-varying geoclimatic metrics applied in the area of ecological niche modeling^35^. They are derived from standard climatic variables such as precipitation and temperature, over time, for a given region. The “dismo” package^36^ in R was used to calculate these. In addition, indices for intradecal climatic systems such as the “El Nino South Oscillation” (ENSO) and the “North Atlantic Oscillation” (NAO) were obtained using the “rSOI”^37^ package in R. To account for time-lag effects, all climatic variables were aggregated into four quarters as done in similarly scoped studies^38^. This excluded the Bioclimatic variables which are defined annually.

Environmental and host data was obtained directed from the European Environment Agency (EEA)^39^. This included all Corine Land Cover (CLC) and Natura 2000 Bioregions data^40^. Geospatial species distribution data was also obtained from the EEA, by merging the Natura 2000 Habitats and Species tables. This included all species, to allow for potentially unreported hosts. Economic and sociodemographic data was obtained using the “Eurostat” package^41^. The European Center for Disease Control (ECDC) provided mosquito distribution data; this included all reported vectors in the region. West Nile virus case data was also obtained from the ECDC.

WNV case data was quality-controlled to exclude cases likely to be subject to regional variation in reporting or other biases. Excluded were 1) asymptomatic cases, 2) cases with no syndromatic description, and 3) cases acquired outside the reporting district. This is consistent with previous work, in which all but the most severe neuroinvasive manifestations (WNND) were excluded^42^.

This final data set was then divided into a training set (2010-2016) and test/validation set (2018). The statistical effect of large-scale climatic systems and associated downstream covariation may potentially overlap year-to-year. To mitigate, 2017 was omitted from training. Summary statistics and relevant meta-data are provided in *Supplementary Table 1*.

### Synthetic data generation

The XGBoost package in R^24^ was used to generate the predictive model from which the SHAP meta-model would be generated. For classification tree models, iteration limits are commonly set so as to avoid over-fitting. Cross-validation was therefore applied to determine this limit: the training set was split according to year and a model was trained using six of the seven years, and the seventh (“hold out”) year was then used to evaluate performance. This process was repeated seven times, using each of the included years once as the hold out set. The cross-validation process diagnostically evaluates the modelling process and determines optimal hyperparameter settings, including the optimal number of iterations. This iteration limit was therefore applied to a train a model using the entire training data. An additional criteria was set to stop model training after 10 iterations with no reduction in log-loss error. Covariates found to be insignificant with respect to three importance criteria – marginal contribution to predictive output (“gain”), internal predictive prevalence (“frequency”), and case-wise predictive relevance (“cover”) – were dropped and the model was run once again with the selected, significant feature set.

XGBoost is notoriously sensitive to hyperparameter settings^43^. Hyperparameters were therefore set to align with traditional statistical standards: 1) 95% was set as the internal discriminatory threshold, 2) each submodel was limited to half of the total feature space, and 3) maximum tree size was set to twice the number of included feature classes (i.e., 16). Consistent with guidance put forth in the seminal paper by Elith J. et al (2008), step size was reduced to 0.03 (10% of the default) to ensure possible feature interactions were exhaustively assessed. No further “tuning” of hyperparameters was attempted.

The “SHapley Additive exPlanations” (SHAP) engine is an AI-driven deductive framework for imputing local feature effects^23^. The underlying algorithm is derived from a military intelligence application based on cooperative game theory. This algorithm determines individual contributions for group outcomes and has been mathematically proven to be optimal with respect to this task^25^. The SHAP implementation is applied to an XGBoost model to determine the degree to which individual features influence the computed event probability for each case. The output of this process is a meta-model, consisting of an effect matrix dimensionally identical to the original, nominal value data (matrix): one complete set of feature effect estimations for each case. Each term reflects the local change in log relative risk associated with a given feature with respect to the global, baseline probability of event. This global baseline risk is computed along with the effect matrix and is the same for all cases. This transformation effectively recenters the predictive output at zero, implying a predictive threshold equivalent to the computed baseline probability. Values greater than zero indicate increased relative risk of event and *vice versa*. This transformation provides for several useful analytical modalities that are presented later and in the main text. The inputs for this process are the computed XGBoost model and the original, nominal value data (set).

The SHAP engine was therefore applied to this model to generate a SHAP meta-model for the 2018 validation period. The choice of classification threshold directly affects predictive performance and several methods exist to optimize^44^. However, as with manual or automated tuning of hyperparameters in XGBoost, the effect of such methods on out-of-sample performance is often unpredictable^45^. Out-of-sample performance was therefore evaluated at the threshold inherent to the SHAP transformation – zero. In-sample performance was likewise estimated using the 2010-2016 training set. For each, the following metrics were assessed: 1) AUC, 2) sensitivity, 3) specificity, and 4) balanced accuracy. AUC is the probability that a randomly selected positive case will be assigned a higher probability than a randomly selected negative case. It indicates how well the model discriminates between positive and negative classes irrespective of classification threshold. The remaining three metrics are threshold-dependent and vary accordingly. Sensitivity, also referred to as true positive rate, measures the probability of detecting event occurrence. Specificity, also referred to true negative rate, measures the probability of correctly identifying non-occurrence. Balanced accuracy is the average of the prior two metrics; it measures the probability of detection assuming negative and positives classes are equally represented in the data set. In the present work, positive classes represent only 0.05% of the total sample. A model that optimizes balanced accuracy irrespective of threshold is therefore preferable. The SHAP-implied classification threshold was also applied to the original XGBoost model, on a diagnostic basis, to confirm predictive equivalence.

Correlation analysis, logistic regression, and linear regression were used for validation. Pearson’s correlations were applied to test for linear association between the SHAP effect matrix and original, nominal feature data. Features were then ranked according to mean absolute SHAP effect and added in step-wise fashion to a logistic regression model. This was done for both the original, nominal value matrix and SHAP effect matrix. The predictive objective was binary WNV presence. For each step, pseudo R-squared was computed and compared, for p=1 through p=50 features. Features were subsequently grouped according to feature class and similarly analyzed, for p=1 through p=30 features, for both data sets. Each feature class set was subsequently aggregated and added to a linear regression model, with actual case count as the predictive objective. Goodness of fit was obtained along with parameter estimates for each aggregate.

The “XGBoost”^46^ package in R was used for generation of the initial XGBoost models as well as the SHAP meta-models. Linear and logistic regression were conducted using the built-in R function “lm”. The “ggplot2”^47^ package in R was used for visual summarization and the “tmap”^48^ package in R was used for geospatial mapping. Geospatial polygons were obtained from the Statistical Office of the European Commission using the R package, “eurostat”^41^. All computations as well as as necessary data processing were conducted in R, using the “data.table”^49^ package.

## Data Records

The 2018 data referenced in this work is freely available on Zenodo (Gayle 2020b)^50^. This official repository contains 1) the nominal value predictor data, and 2) the analogous SHAP-derived effect estimates. This data is provided at NUTS3 resolution and is accordingly geotagged. This data is directly readable into R as a geospatial “simple features” data frame.

The code to recreate this map, along with additional data and risk maps, will be made available online via Github (https://github.com/aruberuku/EU2018WNV/) with an interactive version soon to follow. The original training data is available upon request and will be made available online at a later data. Asset data used are publicly readable and directly available from the original sources (see *Methods)*. Users are advised to check the Github repository regularly. This product is still in its early phases but plans are in place to incorporate new analyses and data products, and to expand the methodology as needed. A complete data compendium is provided as *Supplemental Table 1:* a) provides a table of data labels with human-readable names and feature-class groupings, b) provides summary statistics, and c) provides sources and definitions for each included feature.

## Technical Validation

Out-of-sample performance for the underlying XGBoost model approached 99% (96% – 99% AUC), which far exceeds that of any similarly-scoped model. Sensitivity, specificity, and balanced accuracy were 0.89, 0.96, and 0.92 respectively at a SHAP-implied classification threshold of 0.0062. Without further validation, the demonstrated performance alone would imply an underlying determinant structure with a high degree of empirical validity. The following analyses provide further qualification and demonstrate the degree to which the SHAP-derived effect matrix statistically out-performs the original, nominal value data.

### Summary of SHAP specification and estimation process

Many methods exists for deriving statistical effect estimates for multivariate models, and none are considered ideal. Introduced in 1951, the Shapely value estimation heuristic was designed to deliver optimal effect estimates for complex, multivariate systems with respect to a shared outcome. Shapely values represent the “expected marginal contribution of [a given feature] to any random coalition” and have been mathematically demonstrated to be optimal with respect to this task^51^. Shapely values are heuristically determined according to the following axioms:

**Table 1.**
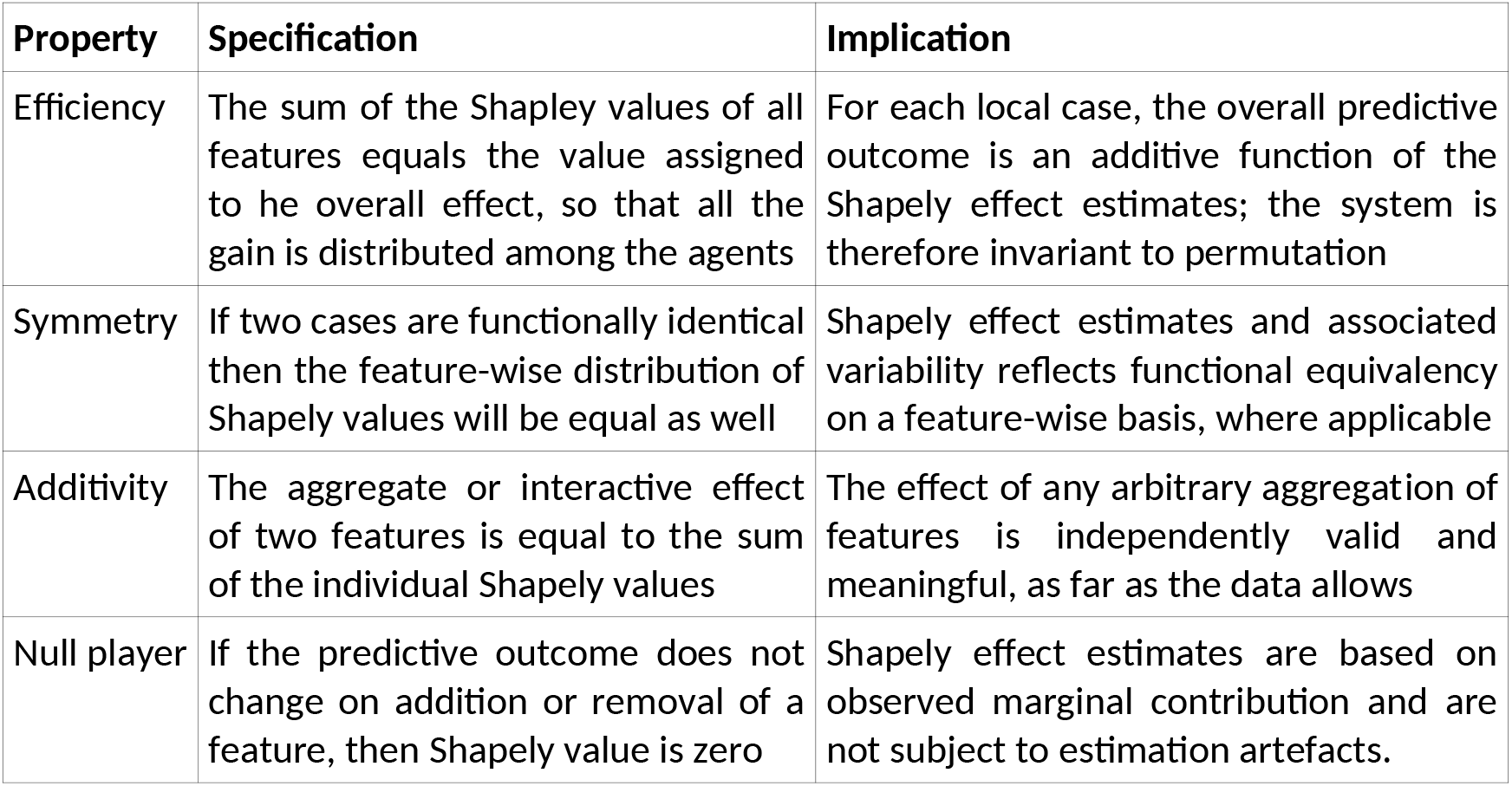
Heuristic axioms applied to generate Shapley value. Likelihood maximization is applied based on these axioms to deductively impute independent feature effects. Simultaneous satisfaction of these conditions yields effect estimations that are proven to be “optimal”.

| Property | Specification | Implication |
| --- | --- | --- |
| Efficiency | The sum of the Shapley values of all features equals the value assigned to the overall effect, so that all the gain is distributed among the agents | For each local case, the overall predictive outcome is an additive function of the Shapely effect estimates; the system is therefore invariant to permutation |
| Symmetry | If two cases are functionally identical then the feature-wise distribution of Shapely values will be equal as well | Shapely effect estimates and associated variability reflects functional equivalency on a feature-wise basis, where applicable |
| Additivity | The aggregate or interactive effect of two features is equal to the sum of the individual Shapely values | The effect of any arbitrary aggregation of features is independently valid and meaningful, as far as the data allows |
| Null player | If the predictive outcome does not change on addition or removal of a feature, then Shapely value is zero | Shapely effect estimates are based on observed marginal contribution and are not subject to estimation artefacts. |

The result of applying this heuristic are local feature effect predictions that are independent individually and on the aggregate. The efficiency and additivity properties therefore allow for meaningful decomposition of the SHAP effect matrix on a case- and feature-wise basis. The symmetry and null player properties furthermore imply that any statistical associations between features exposed by multivariate analyses will be meaningful with respect to any empirically observable relationships between said features and the predictive objective.

### SHAP effect matrix is statistically robust and empirically sound

As implemented in SHAP, local effects are deductively imputed based on the marginal contribution determined via reanalysis of the decision tree ensemble from which the original XGBoost predictions were derived. In the present model, each XGBoost prediction is backed by thousands of effect estimations across all statistically relevant combinations of features. Each local effect estimation is therefore supported by a broad range of experimental configurations, with feature value combinations and predictive output that is unique for each local case. One practical result is that the imputed effects for a given feature will vary independently, on a case-wise basis, even if the original, nominal input values are the same. Lower resolution data will therefore be projected into the higher resolution predictive space, with an increase in empirical precision. Imputed SHAP effects are therefore both a) highly correlated with the original, nominal value data *(Figure 1a)* and b) overall better predictors than the nominal value data—even when estimated prospectively, on an out-of-sample basis (*Figure 1b)*.

**Legend F1a,b.**
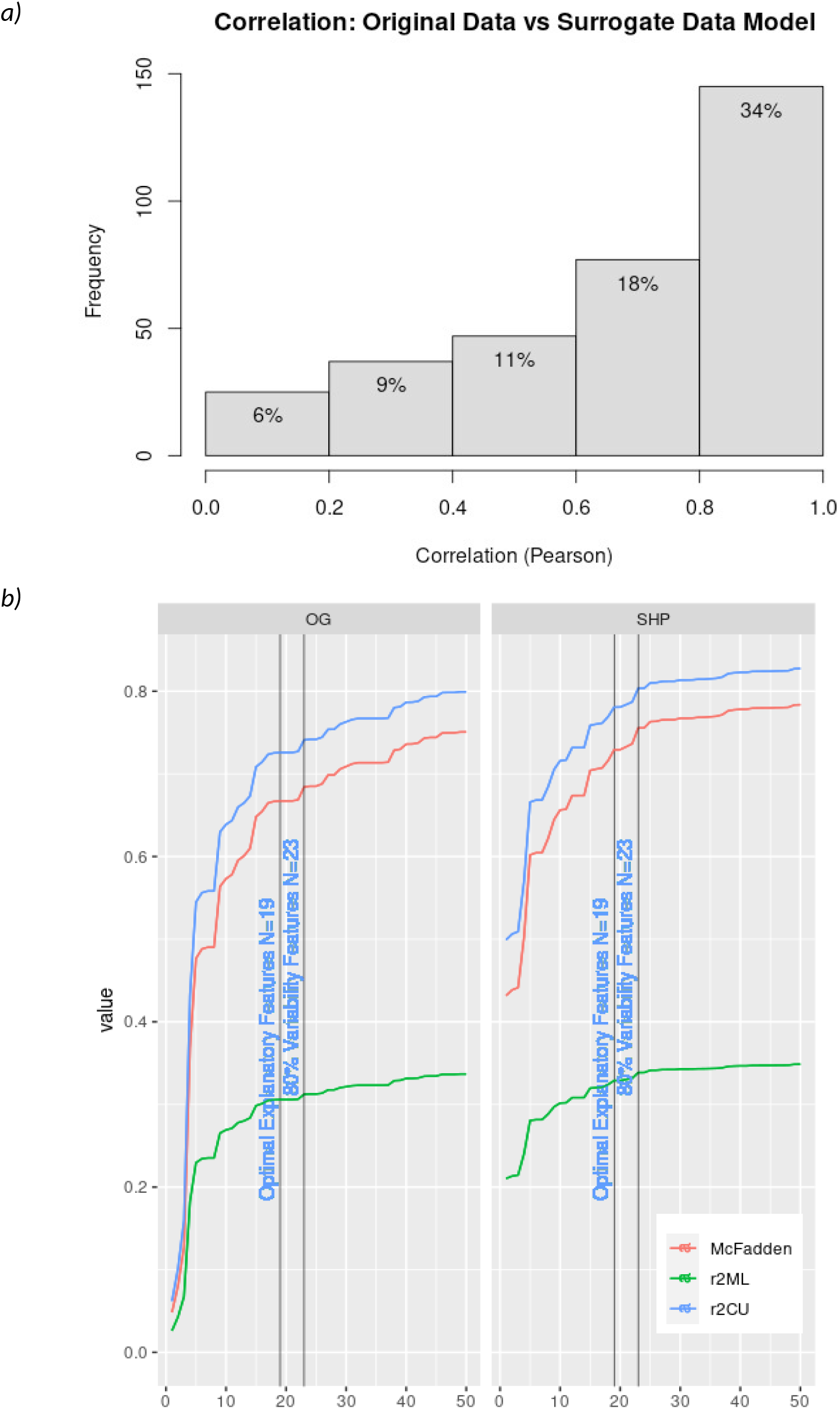
Surrogate data model aligns with but outperforms original data. The surrogate data model is heuristically designed to reproduce the output of the original (XGB) model while estimating the individual, localized effect of each feature. The surrogate data model is therefore dimensionally identical to the original data set but reflects deductively approximated effect estimates. We confirmed a high-degree of statistical correspondence between the two *(Figure 1a*). Stepwise logistic regression *(Figure 1b*) furthermore confirmed that the variability encoded within the surrogate data model aligned better with the original case data, for any given feature set size. This observed difference in predictive performance notably increases as model dimensionality is reduced. This finding is consistent across under various scenarios of model decomposition – e.g, by feature class.

Recall that all feature-wise constellations are specified to be equally and independently valid with respect to the predictive outcome. In the present case, six distinct feature classes were included as predictors and logistic regression revealed each set to independently meaningful with respect to outbreak risk prediction *(Figure 2a)*. SHAP effect estimates were furthermore consistently – and in some cases, substantially – found to outperform the original, nominal value data, particularly at lower feature set sizes. Validity was furthermore qualitatively confirmed via geospatial visualization: each domain-specific map independently reflected meaningful, domain-specific knowledge relevant to local outbreak risk (*Figure 2b)*. Regression analysis furthermore confirmed both validity and statistical independence of domain-specific aggregations even when applied to alternative outbreak criteria – in this case, total case count per region (*Table* 2). This is a considerably more sensitive predictive objective and degree of performance demonstrated speaks to the empirical robustness of the SHAP estimates. Empirical validity of the SHAP-derived outbreak risk map and implications with respect to local and cross-scale determinant relationships are treated in two forthcoming publications. Others are welcome to explore and evaluate this data further.

**Legend F2a.**
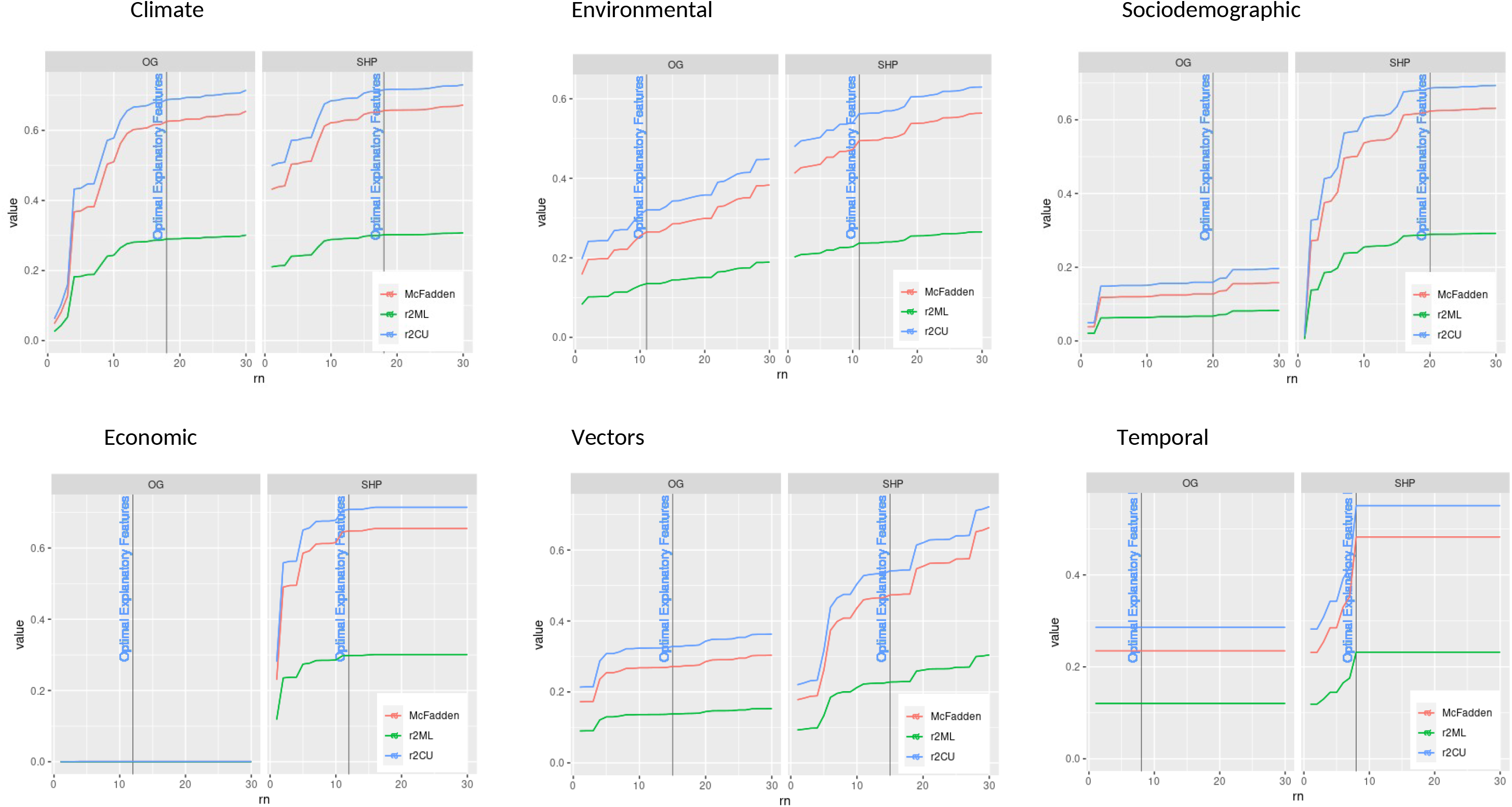
Feature effects predicted by SHAP offer superior statistical robustness. A supplementary analysis was conducted in order to diagnostically assess the properties of the parameters generated by the Shapley transformation. Step-wise logistic regression series were generated for feature class; AUC performance is shown here plotted as a function of the number of included features; hosts showed anomalous results due to limited cases for each included feature and are not shown. The Shapley transformation deductively infers the local effect of each feature with respect to local outcome. These evaluations are internally independent, case-to-case, resulting in a variability profile markedly different from the original data. For example, low-resolution features that are homogenous region to region are still likely to be assigned unique values by the Shapley transformation. The SHAP-derived effect matrix was found to outperform the original, nominal value data in multivariate prediction, even when decomposed according to feature class. The magnitude of performance increase was most pronounced for the feature classes comprised of lower-resolution data. However, even in the case of climate, the benefit of SHAP is undeniable particularly with smaller feature set sizes.

**Legend F2b.**
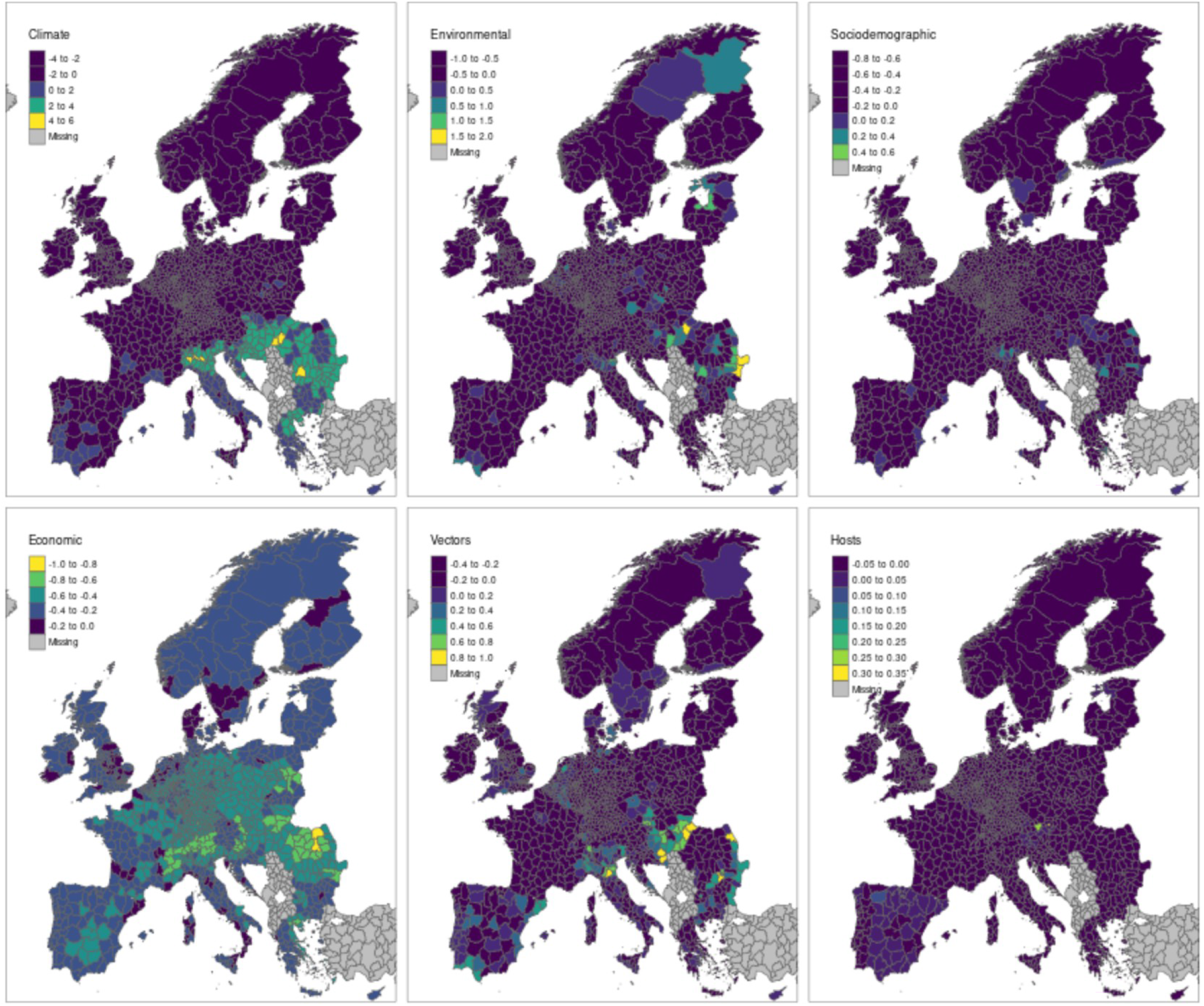
SHAP model decomposition reveals domain-specific patterns of outbreak risk. The ability to arbitrarily decompose a given prediction presents powerful new opportunities. Risk maps were generated according to feature class and rescaled for visual clarity; values are summative with respect to the overall likelihood estimates of West Nile virus outbreak in 2018. The differential effect of the each feature class was found to vary substantially as a function of geography and scale. The effect of climate was confined mostly to the southern latitudes with some notable spread into France and Germany. Risk associated with the environmental feature class had a notable western orientation with homogenous degrees of risk extending from Greece in the south to Sweden in the north. Outbreak risk associated with economic disparities was markedly widespread across, with the Scandinavian countries and UK being among the most notable exceptions. Presence and diversity of potential hosts contributed most notably to outbreak risk in Spain. As for known and potential mosquito vectors, our results showed a far more sporadic pattern of risk. In addition to being substantial in the WNV endemic regions of Italy and Romania and neighboring, vector-associated risk was identified almost uniformly throughout Spain and even into parts of Sweden, Denmark, and nearby Germany.

**Table 2.** Parameter estimates and R-squared results from linear regression. Significant parameters along with goodness of fit metrics confirms empirical robustness of SHAP effect estimates.

|  | Estimate | Std. Error | t-value | Pr(>\|t\|) |  |
| --- | --- | --- | --- | --- | --- |
| **(Intercept)** | 3.51 | 0.45 | 7.75 | 0.00 | *** |
| **Climate** | 0.80 | 0.07 | 12.16 | 0.00 | *** |
| **Economic** | -0.37 | 0.96 | -0.39 | 0.70 |  |
| **Environmental** | 0.84 | 0.31 | 2.72 | 0.01 | ** |
| **Sociodemographic** | 3.40 | 0.89 | 3.81 | 0.00 | *** |
| **Vectors** | 6.51 | 0.77 | 8.47 | 0.00 | *** |
| **Hosts** | -17.11 | 9.69 | -1.77 | 0.08 | . |
| *R*<sup>2</sup>: | 0.230 |  |  |  |  |
| *Adj. R*<sup>2</sup>: | 0.228 |  |  |  |  |

## Usage Notes

**The SHAP effect matrix is dimensionally identical to the original**, nominal value data and is provided here in tabular form. As demonstrated, any analytical steps or methods suitable for the original data would be equally valid with the SHAP effect matrix. Indeed, the SHAP effect matrix is superior in this respect: due to its statistical properties – IID and homogenous with respect to unit, scale, and interpretation – multivariate analyses involving, for example, disparate feature classes and cross-scale effects can be executed without need for methodologically cumbersome preprocessing. Some basic analytical applications are presented in a forthcoming manuscript, available as a pre-print online (Gayle 2020b)^52^. Methods demonstrated include: 1) means testing (student’s t-test) to assess differential drivers of increased outbreak risk in 2018, 2) standardized effect size estimation to “control” for scale-dependent effect variability, 3) and affinity propagation clustering to identify and deterministically enumerate geospatial regions with homogenous outbreak risk profiles.

Others are welcome to explore and evaluate the analytical opportunities represented by this work. Some suggestions might include estimation of the SHAP effect of individual mosquito vectors as a function of the nominal values of various climatic features. Another suggestion might be cluster-based analyses to determine the interactive effects of various combinations of host, vectors, and classes of environment with respect to outbreak risk. Country and region-specific analyses are also possible, along with comparative analyses based on categorical feature range stratifications: for example, regions with high/low population density or high/low proximity to water bodies. In additional, external data could also be married to this data with minimal effort – provided geospatial tags are available. In all cases, we recommend the R platform for ease of combined geospatial and statistical analysis.

## Data Availability

The 2018 data referenced in this work is freely available on Zenodo. This official repository contains 1) the nominal value predictor data, and 2) the analogous SHAP-derived effect estimates. This data is provided at NUTS3 resolution and is accordingly geotagged. This data is directly readable into R as a geospatial "simple features" data frame. 
The code to recreate this map, along with additional data and risk maps, will be made available online via Github (https://github.com/aruberuku/EU2018WNV/) with an interactive version soon to follow. The original training data is available upon request and will be made available online at a later data. Asset data used are publicly readable and directly available from the original sources (see Methods). Users are advised to check the Github repository regularly. This product is still in its early phases but plans are in place to incorporate new analyses and data products, and to expand the methodology as needed. A complete data compendium is provided as Supplemental Table 1: a) provides a table of data labels with human-readable names and feature-class groupings, b) provides summary statistics, and c) provides sources and definitions for each included feature.

https://10.5281/zenodo.3998435

## Code Availability

All programming code necessary to reproduce the map is supplied together with the data (see Data records) and on https://github.com/aruberuku/EU2018WNV/. The Github also hosts several sample maps derived from this data set, including *Figure 2b*.

## Acknowledgements

Funding was received from the European Center for Disease Control and Prevention (contract no ECDC.9504). We would like to thank the European Center for Disease Control for kindly providing funding, case and vector data, and support. And special thanks to the team at Exploratory.io who provided technical guidance and support.

## Author contributions

The authors worked collaboratively and contributed equally to the article.

## Competing interests

The authors declare no competing interests.

## Notes

### Competing Interest Statement

The authors have declared no competing interest.

### Summary of Updates

Complete revision. Supplementary tables added summary statistics and variable definitions.

